# Assessing the effect of global travel and contact reductions to mitigate the COVID-19 pandemic and resurgence

**DOI:** 10.1101/2020.06.17.20133843

**Authors:** Shengjie Lai, Nick W Ruktanonchai, Alessandra Carioli, Corrine W Ruktanonchai, Jessica R Floyd, Olivia Prosper, Chi Zhang, Xiangjun Du, Weizhong Yang, Andrew J Tatem

## Abstract

Travel and physical distancing interventions have been implemented across the World to mitigate the COVID-19 pandemic, but studies are needed to quantify the effectiveness of these measures across regions and time. Timely population mobility data were obtained to measure travel and contact reductions in 135 countries or territories. During the 10 weeks of March 22 – May 30, 2020, domestic travel in study regions has dramatically reduced to a median of 59% (interquartile range [IQR] 43% - 73%) of normal levels seen before the outbreak, with international travel down to 26% (IQR 12% - 35%). If these travel and physical distancing interventions had not been deployed across the World, the cumulative number of cases might have shown a 97-fold (IQR 79 – 116) increase, as of May 31, 2020. However, effectiveness differed by the duration and intensity of interventions and relaxation scenarios, with variations in case severity seen across populations, regions, and seasons.

**One Sentence Summary:** Travel and physical distancing interventions across the World are key to mitigate the COVID-19 pandemic and resurgence.

## Main Text

The COVID-19 pandemic has resulted in 6.3 million confirmed cases and 380 thousand deaths reported across the World as of June 3, 2020, causing an evolving global public health and economic crisis (*1, 2*). As no safe and effective vaccines or pharmaceutical agents are yet known to be available to prevent or treat COVID-19, the medical and public health community are relying on non-pharmaceutical interventions (NPIs) (*3–5*). Travel and physical distancing interventions have been widely implemented across countries by quarantining geographic “hot spots” and minimizing physical contacts between infectors and infectees (*4–7*), aiming to suppress and delay the peak of the first wave in this pandemic, protect healthcare capacity, and reduce the morbidity and mortality caused by COVID-19 (*5, 8–10*).

The implementations of travel reduction and physical distancing measures, together with other interventions, e.g. improved testing, contact tracing and personal behavioral hygiene, are likely to have substantially reduced transmission and flattened epidemic curves across countries (*4, 6, 7, 11*). However, how intensive and effective these travel and social contact interventions across the World are remains unclear, due to the varying durations and intensities of interventions conducted across space and time. Additionally, to minimize the socioeconomic impacts of lockdowns or travel restrictions, plans for relaxing interventions have been proposed and gradually implemented. For example, China seems to have moved past the first wave of the COVID-19 outbreak and lifted travel restrictions that were strictly implemented between late January and early March (*12*), and New Zealand has claimed no community transmission of COVID-19 as their lockdown has been eased (*13*), while travel and social distancing interventions remain in force for many nations across Europe, the Americas and Africa for the time being.

Premature and sudden lifting of interventions could lead to a resurgence and earlier secondary peak (*3, 14, 15*), and an uncontrolled outbreak in one country might introduce transmission in another country (*9*). Certain travel and physical distancing measures, e.g. cancelling mass gatherings, may need to be maintained for a significant period of time to mitigate the risk of resurgence (*16*). No studies have been conducted using quantitative measures of global travel and contact reductions to inform how and when country-based social distancing measures should be implemented or lifted across the globe to mitigate current and future waves of the COVID-19 pandemic in the absence of a vaccine or effective treatment (*14, 15*). To answer these questions, the effectiveness of interventions and potential relaxation strategies across countries should be measured and assessed to guide ongoing and future COVID-19 responses globally.

Anonymized and aggregated human movement data derived from mobile devices have been increasingly used to provide an approximation of population-level mobility and physical contacts during the COVID-19 pandemic (*17–19*). These data can help refine interventions by providing timely information about changes in patterns of human mobility across space and time to support epidemic control strategy design (*20–23*). Here we use timely population movement data to measure the intensity and timing of actual travel across 135 countries or territories under the first wave of the COVID-19 pandemic. A metapopulation transmission model was built to (i) simulate COVID-19 spread across these countries from December 1, 2019 through December 31, 2020, (ii) assess the relative effectiveness of travel and physical distancing interventions that have been in place, and (iii) examine various relaxation strategies. The potential numbers of age-specific severe and critical COVID-19 cases by populations, regions, and seasons were estimated to help guide healthcare resource preparedness.

### Intensity of travel and physical distancing interventions

To quantify the intensity of travel across the World, we used two anonymized and aggregated mobility datasets obtained from Google and Baidu, respectively. The Google COVID-19 Aggregated Mobility Research Dataset contains anonymized mobility flows across 134 countries and territories aggregated over users who have turned on the Location History setting, which is off by default (*24*). This is similar to the data used to show how busy places are in Google Maps — helping identify when a local business tends to be the most crowded. The Google COVID-19 dataset aggregates flows of people from between S2 cells (*25*) from January 5 - May 30, 2020 which is here further aggregated by origin cell and destination country level (See details in Supplementary Materials). Each S2 cell represents a quadrilateral on the surface of the planet and allows for efficient indexing of geographical data. This dataset was analyzed by researchers at the University of Southampton as per the terms of the data sharing agreement. Similar daily population movement data at prefectural level (342 cities) across mainland China were obtained from Baidu location-based services as of May 2, 2020, to define the Chinese mobility and contact rates during the COVID-19 outbreak. The Baidu dataset is publicly available (*26*). To derive country-specific domestic and international mobility, we aggregated the mobility data from S2 cell or city level to country level, as an approximation of country-level travel and physical contacts. To be comparable across countries and time, the flows were further standardized by the ‘normal’ mobility level before the COVID-19 outbreaks or travel and physical distancing interventions (**Fig. 1**; Supplementary Materials).

**Fig. 1.**
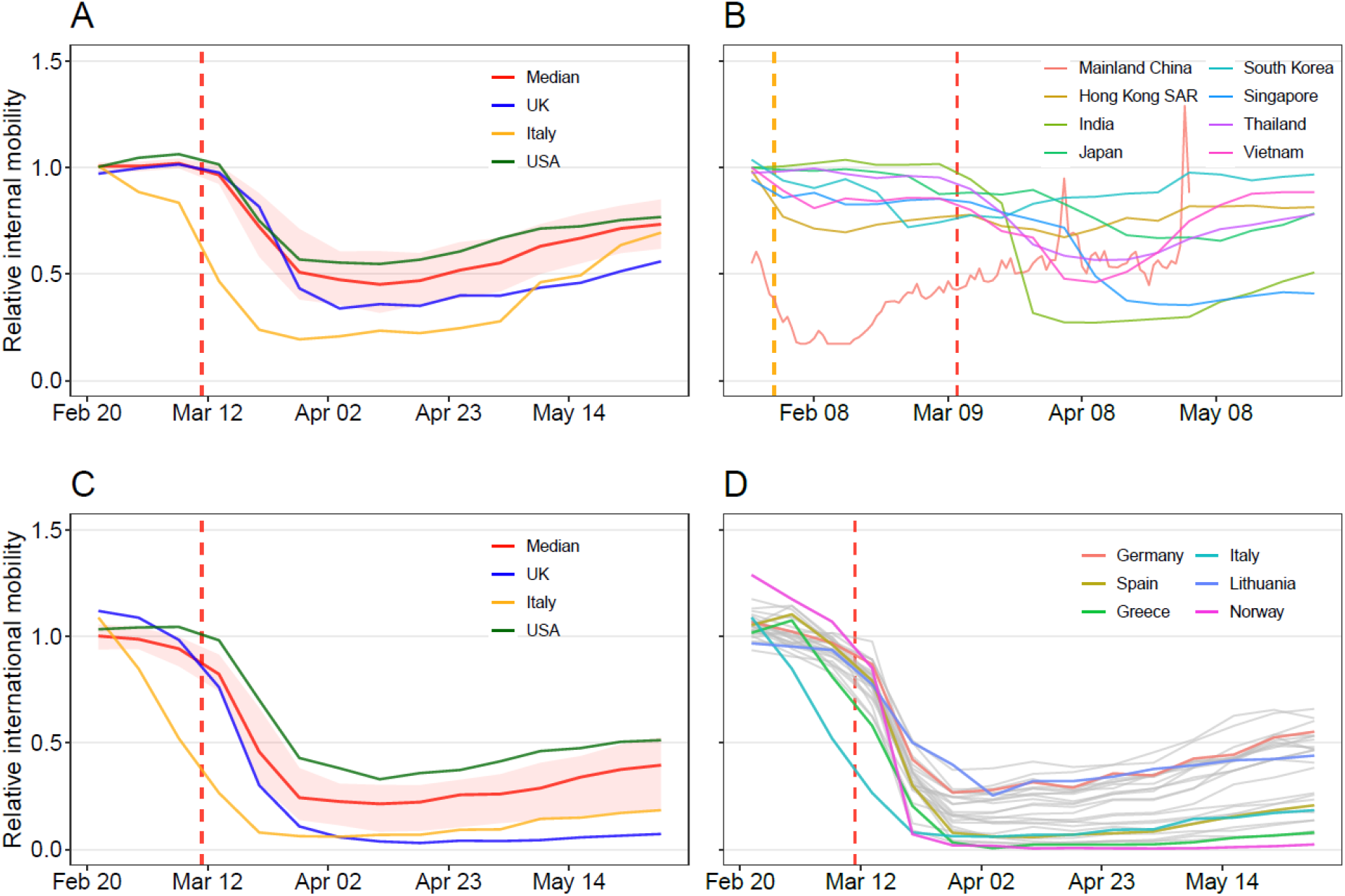
Changing patterns of internal and international population movements across countries or territories, as of May 30, 2020. (**A**) Domestic weekly movements within 127 countries or territories, taking movements on January 5 – February 15, 2020 as a reference. (**B**) Domestic population movements within 8 Asian countries/regions. The daily mobility within mainland China from January 23 to May 2, 2020 was derived from Baidu location-based data, standardized by averaged travel flow on January 5 – 22 before Wuhan’s lockdown on January 23, 2020. All other country or region curves were derived from weekly Google location history data, taking movements on January 5 – 25 as a reference. (**C**) Relative international outflows from 104 countries, with the median and interquartile range provided. (**D**) Relative international outflows from all European countries. The weekly international mobility measures, derived from Google data, took movements on January 5 – February 15 as a reference. The orange and red vertical dashed lines indicate the date of COVID-19 being declared a Public Health Emergency of International Concern and a pandemic by the WHO, respectively. The median (red line) and interquartile range (pink areas) are provided in (**A**) and (**C**), with the curves of Italy, the United Kingdom (UK), and the United States of America (USA) highlighted. Each curve in (**B**) and (**D**) represents the relative travel pattern of a country or territory. SAR: Special Administrative Region.

Population mobility has rapidly declined since mid-March due to the widespread implementation of travel and physical distancing measures. Low levels of movement continued through April across the 135 study countries or territories (**Fig. 1**). During the 10 weeks of March 22 – May 30, 2020, domestic travel decreased to a median of 59% (interquartile range [IQR] 43% - 73%) of levels seen before the interventions, with international travel down to 26% (IQR 12% - 35%) of normal levels. However, the timing and intensity of these reductions in travel and physical contact differed. The reductions appeared earlier in countries that were initially strongly affected by COVID-19, e.g. China, Italy, and in Asian countries or regions that neighbor mainland China (**Fig. 1**), while the decline in African countries occurred later and was less steep with higher residual travel and social contact levels, compared with other countries around the World (**Fig. 2**). The population mobility have gradually resumed in May, with domestic travel back to a median of 69% (interquartile range [IQR] 56% - 80%) of normal levels and international travel recovered to 35% (IQR 15% - 47%) during the 4 weeks from May 3 – 30, 2020 (**Fig. 1C-D**).

**Fig. 2.**
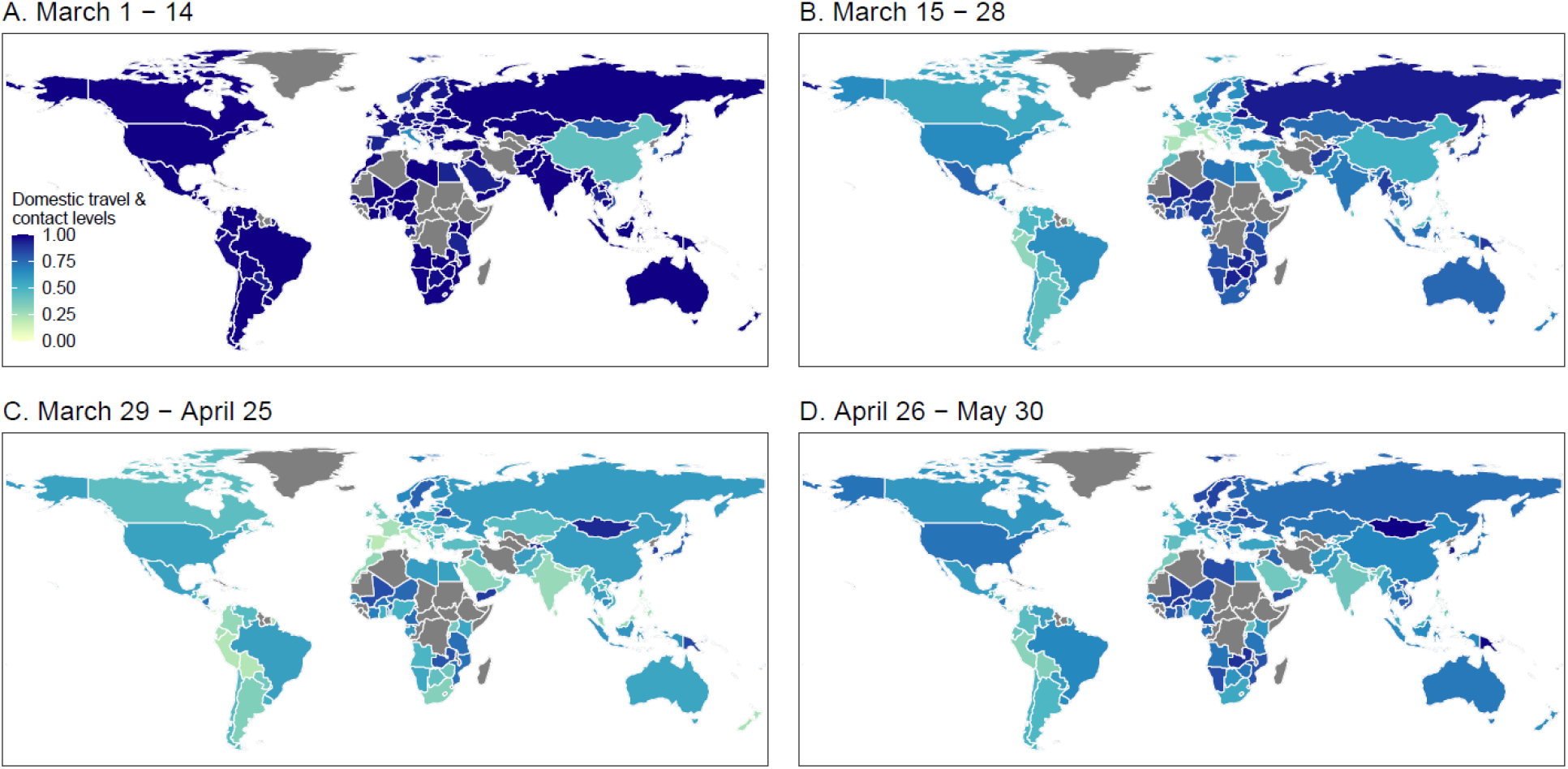
Domestic travel and contact levels by country/territory across 135 countries or territories. (**A**) Population mobility for the two weeks from March 1 - 14, 2020, relative to reference periods. (**B**) Population mobility for the two week of March 15 - 28, 2020. (**C**) Population mobility during the four weeks from March 22 to April 25, 2020. (**D**) Population mobility for the five weeks from April 26 – May 30, 2020. The daily mobility data in mainland China derived from Baidu location-based data as of May 2, 2020 were standardized by the mobility levels on January 5 – 22, 2020. All other 134 countries or territories used Google location history data, with 127 countries or territories taking movements on January 5 – February 15, 2020 as a reference period (**Fig. 1A**), and 7 Asian countries/regions taking movements on January 5 – 25 as a reference (**Fig. 1B**). The administrative boundary maps at 1:110m scales were obtained from the Natural Earth (www.naturalearthdata.com). The regions without data are filled with a grey color.

### Effects of current travel and contact reductions

To quantify the impact of global travel and contact reductions on the COVID-19 pandemic, we used a stochastic susceptible-exposed-infectious-removed (SEIR) modelling framework to simulate the COVID-19 spread in each country or territory. To initially parameterize the model, country-specific effective reproduction numbers (*R*_*e*_) (median 2.4, IQR 2.0 – 2.8) before implementing physical distancing interventions were estimated from daily case counts reported by each country/territory, adjusting for the reporting delays (*27*). We also used the initial epidemiological information (i.e. the incubation period and delays from symptom onset to medical visit/report) estimated for case data at the early stage of the outbreak in Wuhan before the city’s lockdown (*28*). The timings of relevant interventions conducted by each country or territory were derived from a publicly-available dataset of governmental COVID-19 countermeasures (*29*). Finally, the number of cases and the corresponding time-dependent reproduction number in the simulations were calculated under various scenarios of physical distancing interventions and relaxation strategies across regions and time (figs. S1 to S2). The models, parameters, hypotheses, and relevant datasets used are detailed in Supplementary Materials and Data S1 to S2.

As of May 31, 2020, we estimated that there were 15 million (IQR 11 – 20 million) COVID-19 cases under travel and physical distancing interventions deployed across the 135 study countries or territories. These interventions appear to effectively suppress the first wave of pandemic, with 448 million (IQR 365 – 539 million) infections likely prevented in these areas by May 31, 2020. Theoretically, without these interventions, the cumulative number of cases may have shown a 97- fold (IQR 79 – 116) increase as of May 31, 2020, and the peak of the pandemic might occur around July – August, with 51% (IQR 43% - 60%) of the population having been infected across the study regions by the end of 2020. If travel levels were to remain in place through June 30, we estimated that a total of 983 million (IQR: 808 – 1169 million) infections would be prevented by June 30, 2020, with only 20 million (IQR 15 – 27 million) cases developing (**Fig. 3A** and fig. S2).

**Fig. 3.**
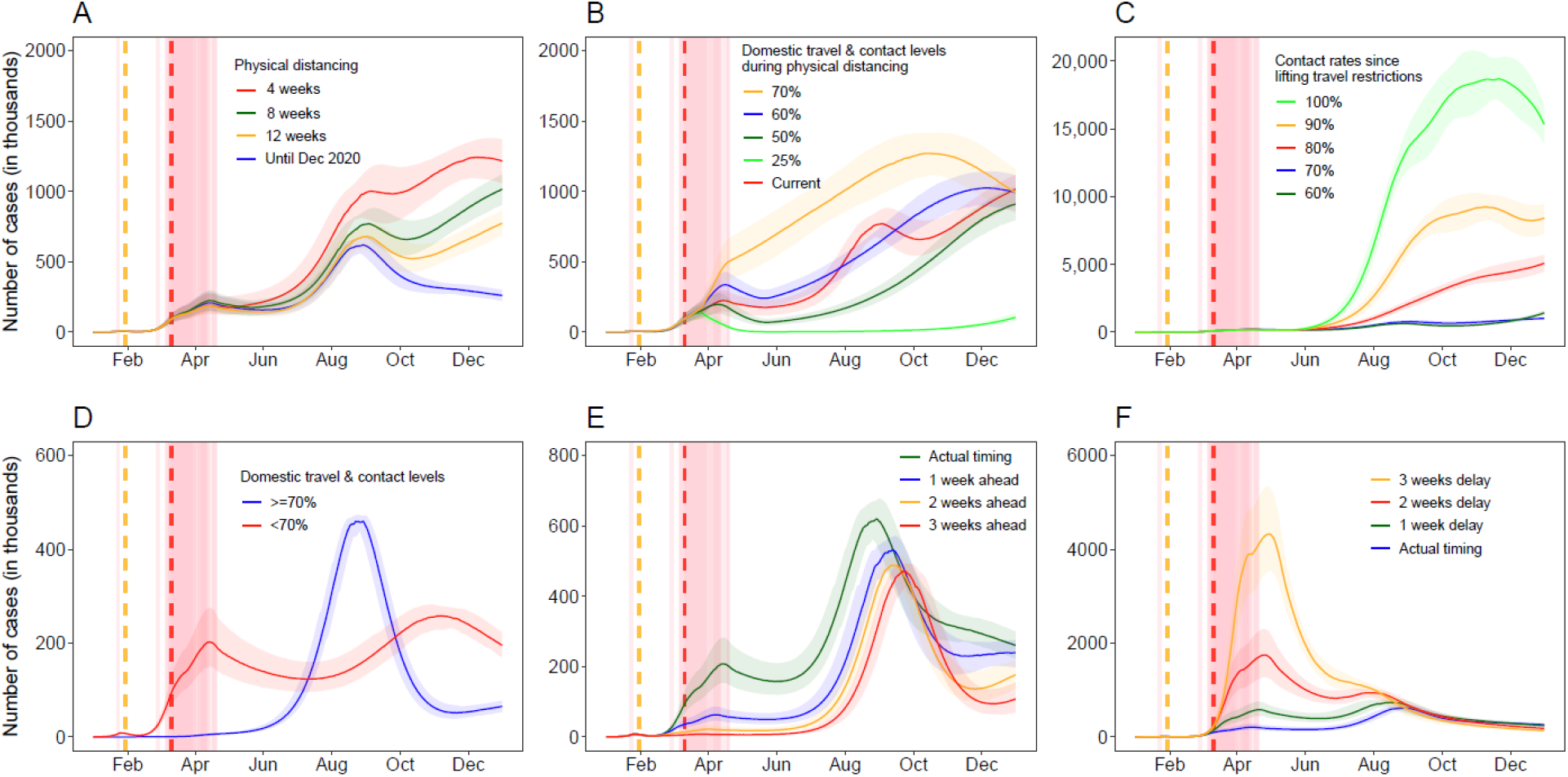
Estimated epicurves of COVID-19 under different intervention scenarios across 135 countries or territories in 2020. (**A**) Implementing travel and physical distancing interventions during various periods. The median and interquartile range of estimates are provided. (**B**) 8-week interventions with various levels of travel and contact rates. In (**A**) and (**B**), the travel and contact levels after relaxing interventions were assumed to be 70% of normal level before outbreaks, if the travel and contact rates in a country or territory were lower than 70%. (**C**) Scenarios of various travel and contact rates after lifting 8-week interventions. (**D**) The estimated epicurves under interventions up until December 31, 2020 based on travel and contact levels by May 2, 2020: 14 countries/territories with travel and contact rates higher than or equal to 70%; 121 countries/territories with the rate less than 70% at any week. (**E**) Estimated epicurves under interventions implemented earlier than actual timing, under the scenario of interventions implemented by December 31, 2020. (**F**) Estimated epicurves under interventions implemented later than actual timing, under the scenario of interventions implemented up until December 31, 2020. The orange and red vertical dashed lines indicate the date of COVID-19 being declared a Public Health Emergency of International Concern and a pandemic by the WHO, respectively. The pink vertical lines indicate the dates of lockdown/physical distancing measures implemented by each country or territory.

The timing of interventions is critical. The COVID-19 outbreak was declared by the World Health Organization (WHO) on January 30, 2020 as a Public Health Emergency of International Concern (*1*). We estimated that, if all travel and physical distancing interventions put in place since February 23, 2020, one month after the lockdown of Wuhan City, had been implemented one, two, or three weeks earlier across the study regions outside of mainland China, the number of COVID- 19 cases by December 31, 2020, would have been dramatically reduced by 33% (IQR 18% – 47%), 50% (38% – 59%), or 58% (48% – 67%), respectively (**Fig. 3E**). If, on the other hand, these interventions had been implemented one, two, or three weeks later than they were, the case count by the end of 2020, would be 1.5-fold (IQR 1.2 – 1.8), 2.5-fold (2.0 – 3.1), or 4.2-fold (3.4 – 5.0) higher, respectively (**Fig. 3F**).

### Impacts of various intervention and relaxation scenarios

Compared with the 4-week travel and physical distancing interventions, the 8-week and 12-week interventions were estimated to further reduce cases by 25% (IQR 20% - 30%) and 39% (IQR 32% - 45%), respectively, by December 31, 2020. However, if the travel and contact levels as of May 2, 2020, could be remained in place through the end of 2020, they would only further reduce the cases by 40% (IQR 33% - 46%) as of December 31, 2020, compared with the 8-week interventions and maintaining travel and contact rates at 70% of their normal levels after relaxing interventions (**Fig. 3A** and fig. S3). If a strict 8-week intervention could be in place across all regions in which there was only 25% of normal travel and contact rates, COVID-19 could be significantly suppressed at a relatively low level of daily new cases (median 4,155, IQR 2,555 – 7,364) from May through September 2020, without a resurgence before October (**Fig. 3B**).

We further assessed the potential effects of various travel and contact rates after easing interventions. We found that, relaxing the interventions would result in an increase in the number of cases, and a complete cancellation of travel and physical distancing interventions will lead to a rapid resurgence of COVID-19 (**Fig. 3C**). If the physical distancing intensity were maintained at 70% of normal levels or lower after relaxing interventions, countries might significantly delay the next wave and reduce its peak. However, due to the heterogeneity of intensities and extents in interventions among countries, a relatively high proportion of populations (median 14%, IQR 11% - 16%) might be infected by the end of 2020 in those countries with weak travel and physical distancing interventions, with only 0.9% (IQR 0.7% - 1.1%) population potentially infected in other countries with more intensive measures (**Fig. 3D**). These differences in interventions would result in temporal and spatial heterogeneity of COVID-19 across the World (**Fig. 4**).

**Fig. 4.**
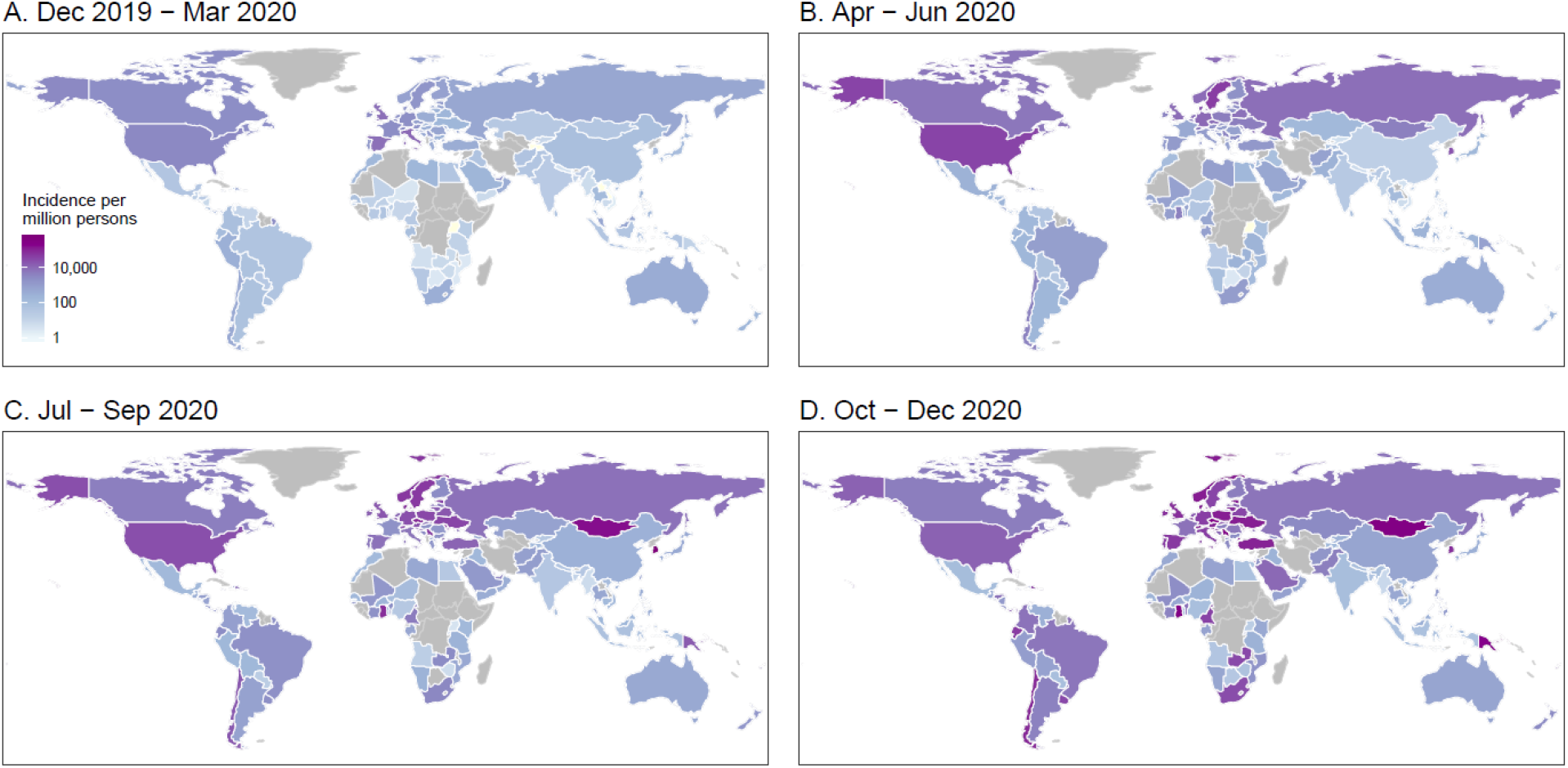
Estimated incidence of COVID-19 cases by season and country/territory (n=135), under 8-week travel and physical distancing interventions. (**A**) Estimated incidence in December 2019 – March 2020. (**B**) Estimated incidence in April – June 2020. (**C**) Estimated incidence in July– September 2020. (**D**) Estimated incidence in October– December 2020. The estimated incidence was standardized by the population counts of each country/territory in 2020. If the travel and contact rates in a country or territory were lower than 70% of normal levels before outbreaks, the rates were assumed to return to 70% after relaxing interventions. The administrative boundary maps at 1:110m scales were obtained from the Natural Earth (www.naturalearthdata.com). The areas without mobile phone data and estimates are filled with a grey color.

To further understand the changing demands on health care from COVID-19, especially utilization of intensive care unit (ICU) beds, we estimated the number of severe and critical cases that would occur across regions and seasons, based on our simulations of COVID-19 spread under the 8-week travel and physical distancing interventions, the age structure of the population in each country/territory in 2020 (37), and the age-specific severity risk of confirmed COVID-19 cases reported from Wuhan (29) (See details in Supplementary Materials). We estimated that a total of 0.9 million (IQR 0.6 – 1.2 million) cases as of March 31, 2020, might have progressed to severe and critical disease, with a potential cumulative number of 33 million (IQR 28 – 39 million) severe and critical cases developing by the end of 2020. Substantial variations in severe and critical infections would be seen across populations, continents, income groups, and seasons (**Fig. 5** and figs. S4 to S6).

**Fig. 5.**
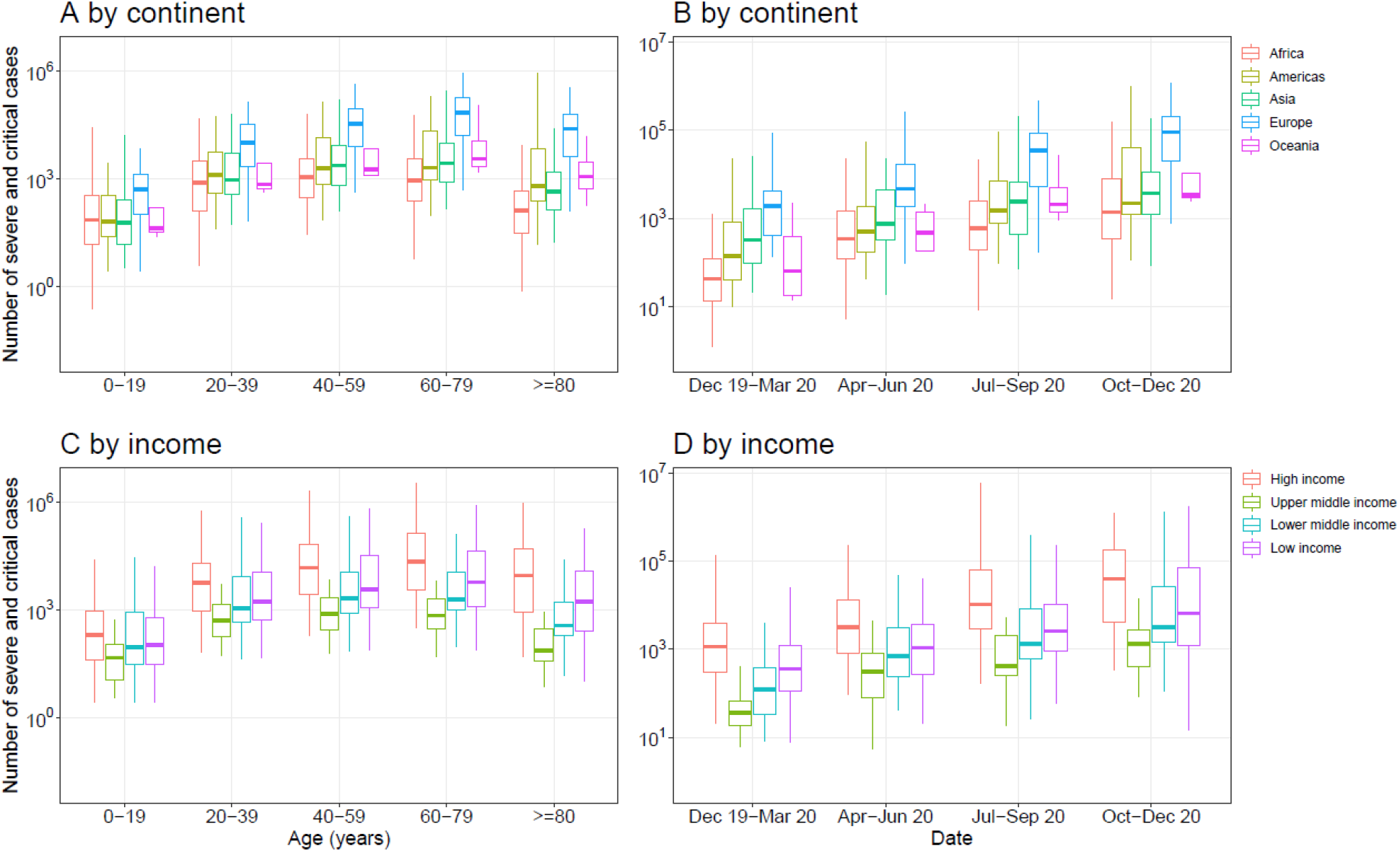
Estimates of severe and critical COVID-19 cases in 135 countries or territories. (**A**) Estimates by age and continent. (**B**) Estimates by season and continent. (**C**) Estimates by age and income classification of each country or territory. (**D**) Estimates by season and income classification of each country or territory. The estimates were based on the scenario of 8-week travel and physical distancing interventions. If the travel and contact rates in a country or territory were lower than 70% of normal levels before outbreaks, the rates were assumed to return to 70% after relaxing interventions. The estimates of severe and critical infections were preliminarily based on the age structures of populations in each country/territory in 2020 (*41*) and the age-specific severity risk of confirmed COVID-19 cases reported from Wuhan over the outbreak (*42*)

We validated our model and outputs using reported case numbers across regions and periods (fig. S7), with a series of sensitivity analyses conducted to help better understand the effectiveness of travel and physical distancing interventions under various settings (figs. S8 to S15). The overall correlation between the number of estimated cases and the reported number by country or territory, as of May 25, 2020, was significant (p<0.001, *R*^2^=0.73), with high correlations also found before (p<0.001, *R*^2^ =0.83) and since (p<0.001, *R*^2^ =0.72) the implementation of travel and physical distancing interventions, respectively. The estimated epicurves of the first wave in this pandemic were also consistent with reported data (p<0.001, *R*^2^=0.91) (fig. S7D).

## Discussion

The COVID-19 pandemic has led to the implementation of unprecedented travel restrictions and physical distancing interventions around the World in 2020. Using near real-time aggregated population mobility data, derived from time- and space-explicit mobile phone data, our study quantified the role of travel and contact changes in mitigating the pandemic across multiple countries. We found that since mid-March, global population movements have dramatically declined and remained at low levels in April, but gradually recovered in May. These multi-nation, aggressive, and continuous measures have played a significant role in suppressing and containing the first wave of the pandemic, and likely prevented a large number of cases, in turn alleviating the pressure on medical and public health services in areas where COVID-19 has spread in the community. Ultimately, these interventions have likely also helped delay subsequent waves of the pandemic, buying time for global preparation and response, and increasing the potential for the development of vaccines and therapeutics that could be used in later stages.

The effectiveness of travel and physical distancing interventions in slowing down COVID-19 transmission, however, hinges on the reduction in the number of contacts between infected individuals and healthy individuals, and between population groups with high rates of transmission and population groups with no or a low level of transmission (*5*). The timing and intensity of physical distancing interventions in countries were not fully synchronized, and the ability to and strategies for containing or mitigating the COVID-19 outbreak also differed. Therefore, the effects of the interventions manifest differently across regions and seasons, especially in low-income countries with weak prevention and control capability (*30*). Current travel restrictions and physical distancing measures have slowed, but not fully contained the COVID-19 pandemic (*31*).

At the end of April 2020, in hopes of reducing the human and economic impacts of social distancing measures, countries are proposed or gradually implementing exit strategies from these interventions and formulating their next pandemic responses. We found, however, that immediately lifting travel and physical distancing measures, would make a second wave of outbreaks inevitable. Similar results have also been seen in previous modelling work (*14, 15*). Moreover, the effectiveness of global travel and contact restrictions varied by the duration of interventions, the intensity of physical distancing, and the travel and contact rates after relaxation of the interventions. Before vaccines or highly effective treatments are available, or widespread herd immunity is achieved, a certain degree of physical distancing, in parallel with early case detection, diagnosis, reporting and (self-) isolation, should be maintained to avoid a rapid resurgence (*4*). In addition, there is currently no evidence that people who recovered from COVID- 19 have immunity to prevent reinfection (*32*). If immunity is not permanent, periodic transmission, e.g. annual cycle, will likely occur (*15*), and physical distancing interventions will again become necessary.

Several limitations in our study should be noted. First, the Google/Baidu data is limited to smartphone users who have opted into relevant product features. These data may not be representative of the population as a whole, and their representativeness may vary by location. Importantly, these limited data are subject to differential privacy algorithms, designed to protect user anonymity and obscure fine detail. Moreover, comparisons across rather than within locations can only be descriptive since regions differ in substantial ways. Second, the accuracy of our model relies on accurate estimates of *R*_*e*_ and other epidemiological parameters derived from reported case data. The quality of reported data and epidemiologic features of COVID-19 likely differs across countries/regions (*33–35*), due to varying case definitions, diagnosis and surveillance capacity, population demographics, and other factors (*36*). Third, we assumed the observed travel and contact reductions have similar effects in minimizing exposure risk of COVID-19 across space and time. The impact of physical distancing might, however, vary between urban and suburban or rural areas with different population densities. Fourth, many other factors may also contribute to COVID-19 spread or mitigation. For example, our simulations did not specify the contributions of pre-symptomatic transmission, presence of other NPIs such as using face masks (*37*), or the potential continuous importations of the virus via international travel, and the seasonal impacts of climatic factors that might have a limited role in the early COVID-19 pandemic (*38*). Fifth, our preliminary estimates of the number of severe and critical cases by age assumed similarity in the severity of cases that were observed in Wuhan, and did not account for the individual characteristics, e.g. comorbidities, and country-specific health care capacity that vary widely across regions and may influence risk of serious disease. Lastly, we did not predict the long-term dynamics of the COVID-19 pandemic over the next few years, or the level of travel and physical distancing interventions that might be required to prevent resurgences, because they depend on a full understanding of the epidemiological features of the disease which are currently lacking.

Viruses do not respect national borders, yet our societies are so deeply interconnected that the actions of one government can have rapid, profound global impacts. Our study serves to quantify important metrics of travel and physical distancing interventions, suggesting their potential effectiveness across the globe to improve international strategies and guiding national, regional and global future responses to prioritize limited resources and strengthen healthcare capacity. Countries vary widely in terms of their ability to prevent, detect, and respond to outbreaks (*39*), and many low and middle income might not be able to provide sufficient access to health care resources in the face of rapid spread of COVID-19 (*40*). For the time being, therefore, travel and physical distancing measures remain still critical tools in mitigating the impacts of the COVID-19 pandemic, and some levels of interventions may be required for considerably longer to prevent a rapid resurgence. Given the improving access of timely anonymized population movement data for supporting COVID-19 mitigation across the World (*24*), the potential exists to monitor and assess the effectiveness of travel and physical distancing interventions to inform strategies against future waves of the COVID-19 pandemic.

## Data Availability

Code for the model simulations is available at the following GitHub repository: https://github.com/wpgp/BEARmod. The data on COVID-19 cases and interventions reported by country are available from the data sources listed in Supplementary Materials. The parameters and population data for running simulations and estimating the severity are listed in Supplementary Data S1 to S2. The population movement data obtained from Baidu are available at: https://qianxi.baidu.com/. The Google COVID-19 Aggregated Mobility Research Dataset used for this study is available with permission of Google, LLC.

## Acknowledgments

The authors would like to acknowledge Google and Baidu for sharing population movement data, and we also thank Yanyan Zhu and Shuhao Lai for supporting data analyses.

## Author contributions

S.L. designed the research. S.L., N.W.R., and O.P. built the model. S.L. ran simulations and carried out analyses. A.C., C.W.R., and J.R.F provided technical support. N.W.R, A.C., C.W.R., and J.R.F helped with data curation. A.C. collated the age-structure data of populations. C.Z. and X.D. collated Baidu mobility aggregated dataset. O.P., C.Z., X.D., and W.Y. did not access to the Google data used in this study. S.L. wrote the first draft of manuscript. S.L., N.W.R., A.C., C.W.R., J.R.F., O.P., C.Z., X.D., W.Y., and A.J.T. commented on and edited the manuscript.

## Data and materials availability

Code for the model simulations is available at the following GitHub repository: https://github.com/wpgp/BEARmod. The data on COVID-19 cases and interventions reported by country are available from the data sources listed in Supplementary Materials. The parameters and population data for running simulations and estimating the severity are listed in Supplementary Data S1 to S2. The population movement data obtained from Baidu are publicly available online at: https://qianxi.baidu.com/. The Google COVID-19 Aggregated Mobility Research Dataset used for this study is available with permission of Google, LLC.

## Ethical approval

Ethical clearance for collecting and using secondary population mobility data in this study was granted by the institutional review board of the University of Southampton (No. 48002). All data were supplied and analyzed in an anonymous format, without access to personal identifying information.

## Supplementary Materials

Materials and Methods Figures S1-S15

Data S1 to S2

